# COVID-19 pandemic in Djibouti: epidemiology and the response strategy followed to contain the virus during the first two months, 17 March to 16 May 2020

**DOI:** 10.1101/2020.08.03.20167692

**Authors:** Mohamed Elhakim, Saleh Banoita Tourab, Ahmed Zouiten

**Affiliations:** Technical Officer, World Health Organization, WHO Representative’s Office, Djibouti-ville, Republic of Djibouti; Executive Secretary, Ministry of Health of Djibouti, Djibouti-ville, Republic of Djibouti; WHO Representative, World Health Organization, WHO Representative’s Office, Djibouti-ville, Republic of Djibouti

**Keywords:** Coronavirus, COVID-19, Epidemiology, Pandemic, Djibouti

## Abstract

**Background:** First cases of COVID-19 were reported from Wuhan, China, in December 2019, and it progressed rapidly. On 30 January, WHO declared the new disease as a PHEIC, then as a Pandemic on 11 March. By mid-March, the virus spread widely; Djibouti was not spared and was hit by the pandemic with the first case detected on 17 March. Djibouti worked with WHO and other partners to develop a preparedness and response plan, and implemented a series of intervention measures. MoH together with its civilian and military partners, closely followed WHO recommended strategy based on four pillars: testing, isolating, early case management, and contact tracing. From 17 March to 16 May, Djibouti performed the highest per capita tests in Africa and isolated, treated and traced the contacts of each positive case, which allowed for a rapid control of the epidemic.

**Methods:** COVID-19 data included in this study was collected through MoH Djibouti during the period from 17 March to 16 May 2020.

**Results:** A total of 1,401 confirmed cases of COVID-19 were included in the study with 4 related deaths (CFR: 0.3%) and an attack rate of 0.15%. Males represented (68.4%) of the cases, with the age group 31-45 years old (34.2%) as the most affected. Djibouti conducted 17,532 tests, and was considered as a champion for COVID-19 testing in Africa with 18.2 tests per 1000 habitant. All positive cases were isolated, treated and had their contacts traced, which led to early and proactive diagnosis of cases and in turn yielded up to 95-98% asymptomatic cases. Recoveries reached 69% of the infected cases with R0 (0.91). The virus was detected in 4 regions in the country, with the highest percentage in the capital (83%).

**Conclusion:** Djibouti responded to COVID-19 pandemic following an efficient and effective strategy, using a strong collaboration between civilian and military health assets that increased the response capacities of the country. Partnership, coordination, solidarity, proactivity and commitment were the pillars to confront COVID-19 pandemic.

## 1. Introduction

Since the first reports of emergence of the coronavirus disease 2019 (COVID-19) in Wuhan province, China, in December 2019, the disease has progressed very rapidly (1), declared by the World Health Organization (WHO) as Public Health Emergency of International Concern (PHEIC) on 30 January 2020 (2), then as a Pandemic on 11 March 2020 (3). COVID-19 spread widely in few weeks and affected around 215 countries, territories and areas around the globe in the six WHO Regions (4). As of 16 May 2020, 4,525,497 confirmed cases and 307,395 deaths were reported worldwide (4, 5) with a case fatality rate (CFR) of 6.8%. During the past few months, researchers and the global scientific community found answers to many questions related to this emerging disease, yet many questions remain unanswered and require further research and studies.

COVID-19 affected some countries in a more severe course than others (5). As of 16 May 2020, the ten most affected countries with regards to the numbers of cases were the United States of America with 1,409,452 confirmed cases, Russian Federation with 281,752 confirmed cases, The United Kingdom with 240,165 confirmed cases, Spain with 230,698 confirmed cases, Italy with 224,760 confirmed cases, Brazil with 218,223 confirmed cases, Germany with 174,355 confirmed cases, Turkey with 148,067 confirmed cases, France with 140,008 confirmed cases, and Iran (Islamic Republic of) with 118,392 confirmed cases (4, 5).

Although Africa is considered as the most vulnerable continent considering the overall weaker health systems and infrastructure, the African countries were the last ones to be hit by the pandemic (6). Egypt reported the first case of COVID-19 in the African continent on 14 February 2020 (7, 8), since then, the African countries started to report cases with the first case reported from the sub-Saharan Africa, in Nigeria, on 27 February 2020 (9). As of 16 May 2020, the African countries reported 77,937 COVID-19 confirmed cases including 2,608 deaths (10), with a case fatality rate (CFR) of 3.3%.

Djibouti was not spared by the spread of the COVID-19 pandemic. Situated in the horn of Africa, with a surface area of 23,200 square kilometers (2018) (11) and a population of 962,451 (2018) (12), the country’s biggest asset lies in its strategic position at the southern entrance to the Red Sea, forming a bridge between Africa and the Middle East (13). The first case of COVID-19 officially reported in the country was on 17 March 2020 (14). A member of a foreign military contingent, stationed in Djibouti, who had travelled from his country and was diagnosed on arrival to Djibouti, luckily with no contact with the local population and was eventually repatriated back the following day (14).

Since the first case was detected on 17 March up to 16 May 2020, Djibouti reported 1,401 confirmed COVID-19 cases including 4 deaths (15) with a case fatality rate (CFR) of 0.3%. Since the beginning of February, even before the first case was reported, Djibouti worked with WHO and other partners to develop a preparedness and response plan, and the government implemented a series of many intervention measures to prevent and control the virus circulation in the country; this plan was reviewed and updated in March 2020 (16). The plan in Djibouti was developed along the WHO recommendations for testing every suspected case and follow a strict contact tracing method, according to the identified case definition, in order to limit the number of missed COVID-19 positive cases to the minimum. All positive cases detected were isolated in the treatment and isolation centers prepared by the Ministry of Health for this matter (16).

Since March 2020, the government of Djibouti pursued effective and strict policies starting by the closure of land borders, the port and the airport on 18 March 2020 (17). Many other measures have been taken by the government including the closure of all schools and universities on 19 March 2020; the closure of mosques, other places of worship, social gatherings, bars and night clubs, while the public transportation was limited and non-essential staff of the government were placed on administrative leave on 22 March 2020. Finally, on 23 March 2020, a general confinement by district was announced and was quickly introduced in the capital, Djibouti-ville, where almost 70-75% of the population live (18). This confinement was extended by the government up to 17 May 2020 when the country started gradually the de-confinement measures (19).

As the emergence of COVID-19 represented a threat to the global health system, it was important to conduct this study to examine and analyze the epidemiology of the coronavirus disease (COVID-19) in Djibouti, and to explain how successful was the preparedness and response strategy, followed by the civil and military health authorities and their partners in Djibouti, during the first two months of the response to the pandemic in the country from 17 March to 16 May 2020.

## 2. Materials and methods

### 2.1. Strategy pillars

The strategy adopted by the Ministry of Health was and its partners was perfectly in line with WHO recommendations based on four pillars: testing, isolating, early case management, and contact tracing.

#### Testing

The oropharyngeal and nasopharyngeal samples, as well as, sputum samples were collected from COVID-19 suspected patients and their contacts by a team of paramedics trained for sample collecting at the Hospital Bouffard.

The samples were tested by real time polymerase chain reaction (RT-PCR) machines procured by WHO and also through MoH mechanisms. The reagents used were WHO approved (Sarbecov E-GENE Plus EAV control kits).

#### Isolation and case management

In close collaboration between the civil and military health authorities, a protocol for isolation and case management was developed and implemented first at hospital Bouffard where nearly 50 beds were dedicated to COVID-19 case management. A second center was opened in Arta region using the premises of a hospital used by the social security department (16).

#### Contact Tracing

The Police and Gendarmerie’s medical services were in charge of tracing, quarantining, testing and isolating all the close contacts of each confirmed case.

### 2.2. Study population

All coronavirus disease (COVID-19) laboratory-confirmed cases reported officially by the Ministry of Health of Djibouti during the study period. A case definition for the COVID-19 was identified by the Ministry of Health of Djibouti and it was in line with the WHO case definition during the period of the study.

### 2.3. Case definition used for COVID-19 patients and contacts by MoH of Djibouti (16)

***Suspected case:*** 1) a patient with acute respiratory infection (of sudden onset) or at least one of the following symptoms: cough, fever, shortness of breath AND without an etiology explaining the clinical presentation AND with a history of traveling or residence in a country/ region reporting local or community transmission during the 14 days prior to the onset of symptoms; OR 2) a patient with acute respiratory illness AND who has been in close contact with a confirmed case of COVID-19 during the past 14 days prior to the onset of the symptoms; OR 3) a patient with severe acute respiratory infection, fever and at least one sign/ symptom of respiratory illness (e.g. cough, fever, shortness of breath) AND requiring hospitalization (Severe Acute Respiratory Infection - SARI) AND without any etiology that explains the clinical presentation.

***Confirmed case:*** any person, symptomatic or not, with a lab-confirmed specimen by RT-PCR for the infection of COVID-19, the severe acute respiratory syndrome coronavirus 2 (SARS-CoV-2).

***Close contact:*** a person who, during the 24 hours prior to the onset of symptoms of a confirmed case, shared the same living space (e.g. family, roommate) or had direct face-to-face contact within 1 meter or less of the case and/ or for more than 15 minutes, in a discussion; close friends; class or office mates; close to a case in a transport for an extended period of time; a person providing care of a confirmed case or laboratory personnel handling specimens of a confirmed case, in the absence of adequate equipment of protection.

### 2.4. Study duration

Data of COVID-19 patients reported by the Ministry of Health of Djibouti for two months were collected and included in the study. Since the first case of COVID-19 was reported in Djibouti on 17 March 2020 up to 16 May 2020.

### 2.5. Data source

All COVID-19 data used in this study was collected through the epidemiological surveillance system of the Ministry of Health of Djibouti and retrieved through the daily communiqué developed and published by the Ministry of Health of Djibouti (15), as well as, the daily situation report developed by the Ministry of Health in collaboration with WHO Country Office.

### 2.6. Laboratory methods

Since the beginning of the COVID-19 pandemic, the Ministry of Health of Djibouti identified the laboratory of Hospital Bouffard, situated in the capital, Djibouti-ville, as the only laboratory of reference for the testing and confirmation of COVID-19 cases in the country. Military and civilian laboratory technicians worked together under the supervision of a medical biology specialist from the private sector contracted by the MoH.

The swabs, were collected from the COVID-19 suspected patients and from the contacts of COVID-19 positive cases, during the contact tracing procedure, to get tested by RT-PCR following the WHO guidelines for laboratory confirmation of the emerging disease during the study period (20).

### 2.7. Data management

The collected COVID-19 data from Djibouti was revised, tabulated and entered into a PC Excel file (Microsoft Office). Data cleaning and quality checking were performed. The epidemiological and demographic characteristics were summarized in this study. A P-value less than 0.05 was considered statistically significant. Further data analysis was conducted by epidemiological week against the number of COVID-19 positive cases over the study period to show the coronavirus disease peak in the country, as well as, the positivity rate. R naught (R0) was calculated per week and for the whole period of the study.

### 2.8. Ethical approval

This study was conducted using publicly anonymous online available data published by the Ministry of Health of Djibouti; therefore, no ethical approval was needed.

## 3. Results

From 17 March to 16 May 2020, 1,401 positive COVID-19 cases were reported by the Ministry of Health in Djibouti including 4 deaths with a case fatality rate (CFR) of 0.3% and an attack rate of 0.15%. Twenty-eight cases were primary cases and the rest were contacts of positive cases that were found positive during the aggressive and proactive contact tracing exercise. The peak of COVID-19 pandemic in Djibouti started during the first week of April 2020 and the number of cases started to decline during the third week of the same month *(Figure 1)*.

**Figure 1.**
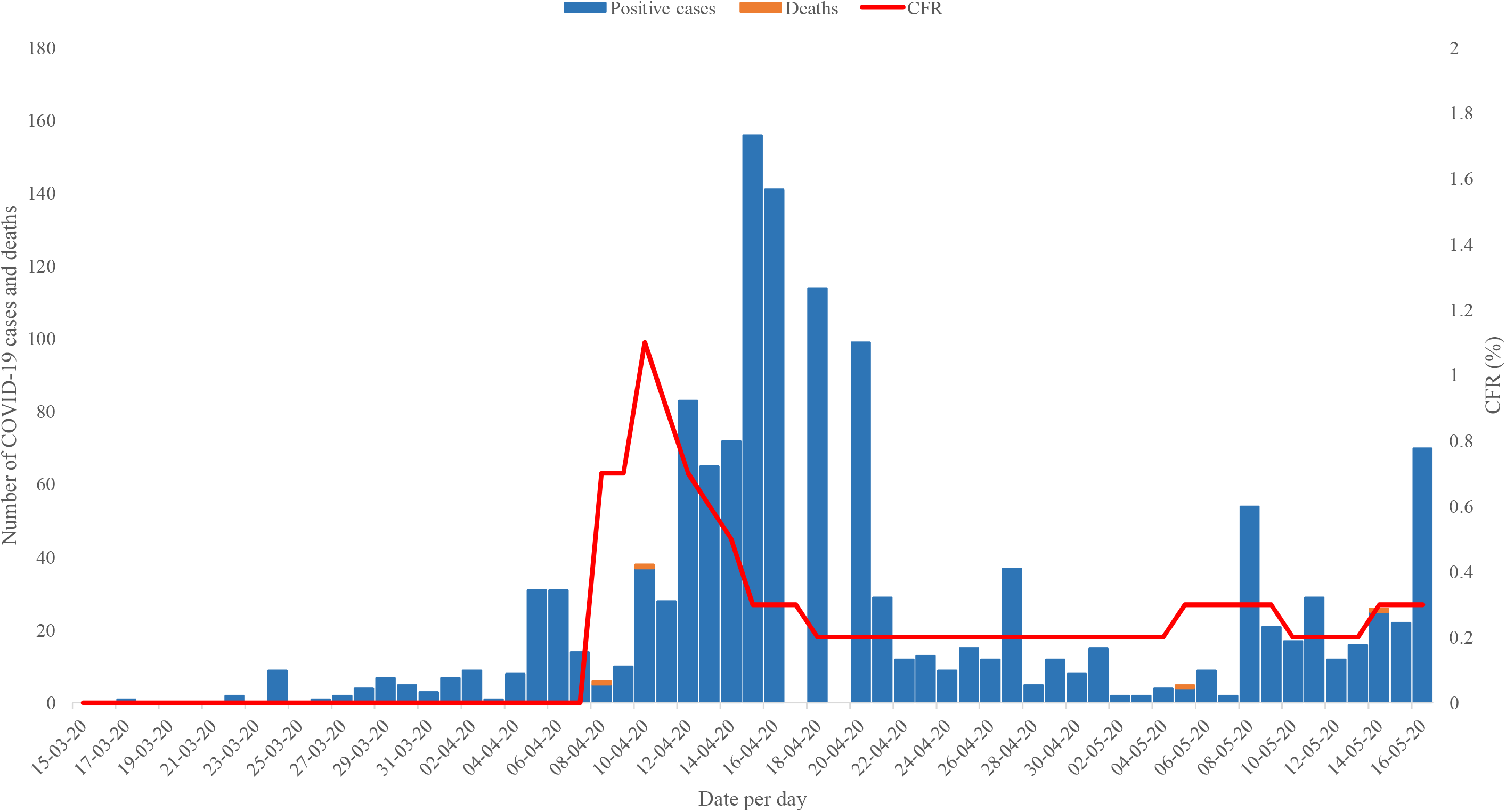
Coronavirus disease (COVID-19) confirmed cases, deaths and case fatality rate (CFR) in Djibouti, from 17 March to 16 May 2020.

The period of this study resides between the epidemiological week number 12 (15-21 March 2020) and the epidemiological week number 20 (10-16 May 2020) (21). *(Figure 2)* shows that the Ministry of Health of Djibouti performed 17,532 tests for COVID-19 during the study period with the epi week 16 (12-18 April 2020) having the highest number of performed tests, 3,693 tests, and also with the highest number of detected positive cases, 631 positive cases detected, with a positivity rate of 17%. The positivity rates since the beginning of the pandemic in Djibouti up to end of epi week 20 varied from 1% in epi week 12 up to 17% in epi week 16, with an overall positivity rate of 8%.

**Figure 2.**
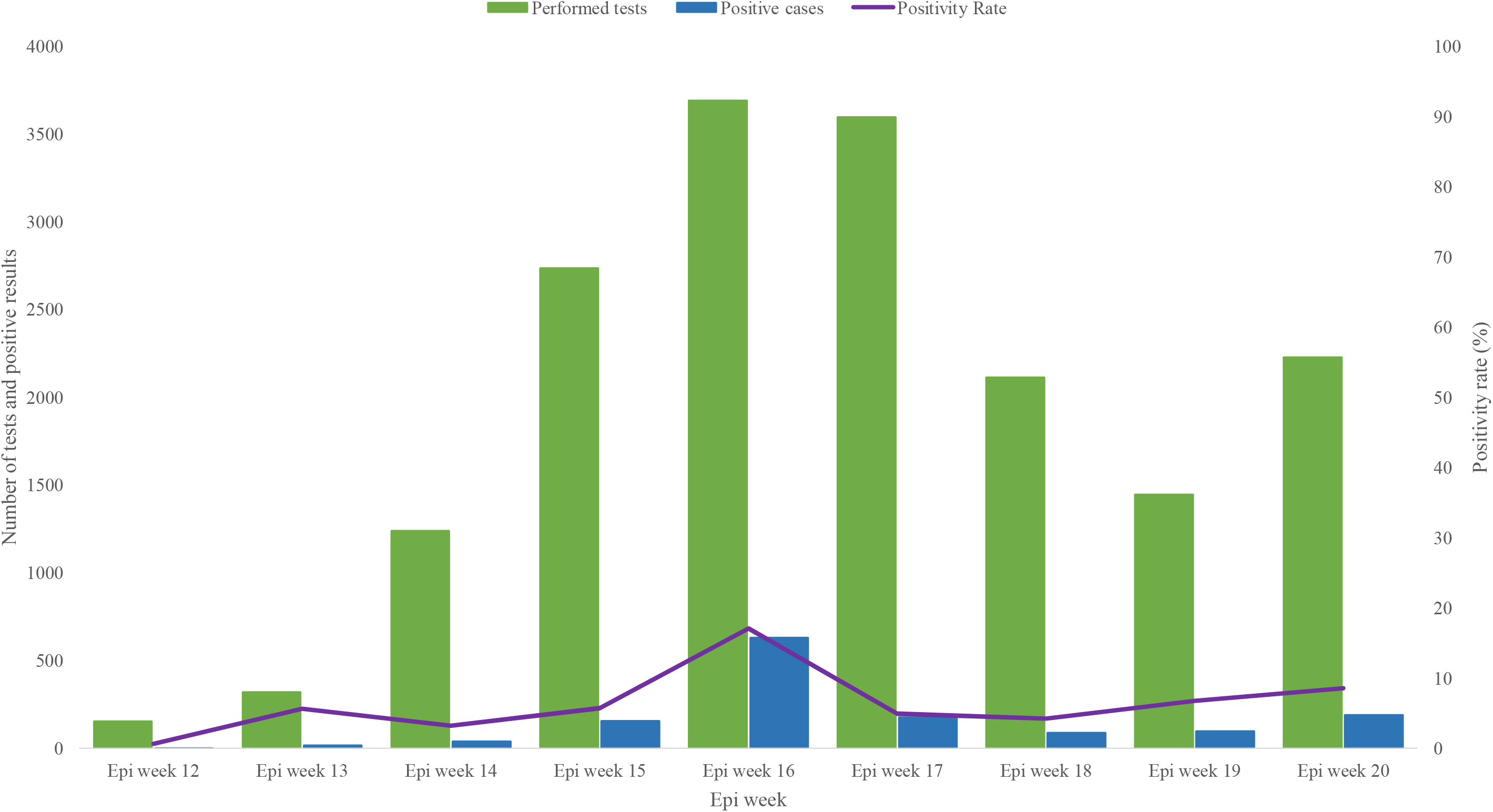
COVID-19 performed tests with the number of positive cases and positivity rate in Djibouti, from Epi week 12 to Epi week 20/ 2020.

The demographic characteristics of COVID-19 cases during the study period in Djibouti shows that male gender dominated the reported cases by 68.4% (958) of the lab-confirmed cases in the country. During the study period, the highest age group affected by the emerging zoonotic virus was the age group 31-45 years old with 34.2% (479) of the cases followed by the age group 16-30 years old with 33.3% (466) of the cases *(Table 1)*.

**Table 1.** Demographic characteristics of the COVID-19 lab-confirmed cases reported in Djibouti, during the first two months of the pandemic, 17 March-16 May 2020.

| Characteristics | Gender (%) |  | Total per age group | P-value |
| --- | --- | --- | --- | --- |
| Age group in years (%) | Male | Female |  |  |
| 0-15 | 60 (4.3) | 20 (1.4) | 80 (5.7) | 0.349653672 |
| 16-30 | 392 (27.9) | 74 (5.3) | 466 (33.3) |  |
| 31-45 | 307 (22.0) | 172 (12.3) | 479 (34.2) |  |
| ≥ 46 | 199 (14.2) | 177 (12.6) | 376 (26.8) |  |
| **Total per gender** | 958 (68.4) | 443 (31.6) | 1401 (100) |  |

The response strategy, based on the four pillars, followed by the Ministry of Health of Djibouti during the study period, along with the treatment protocol agreed upon in the National Action Plan for preparedness and response for COVID -19 (16) showed efficient results during the first two months of the response. This was reflected through a significantly low CFR (0.3%) which was at least 20 times lower that the global CFR reported during the study period (global CFR up to 16 May was 6.8%), as well as, through the high percentage of recoveries reaching 69% (972) of the infected cases in the country. *(Figure 3)* shows the high number of recovered COVID-19 patients during the study period per week with the highest number of recoveries, 365 recoveries, during the epi week 17.

**Figure 3.**
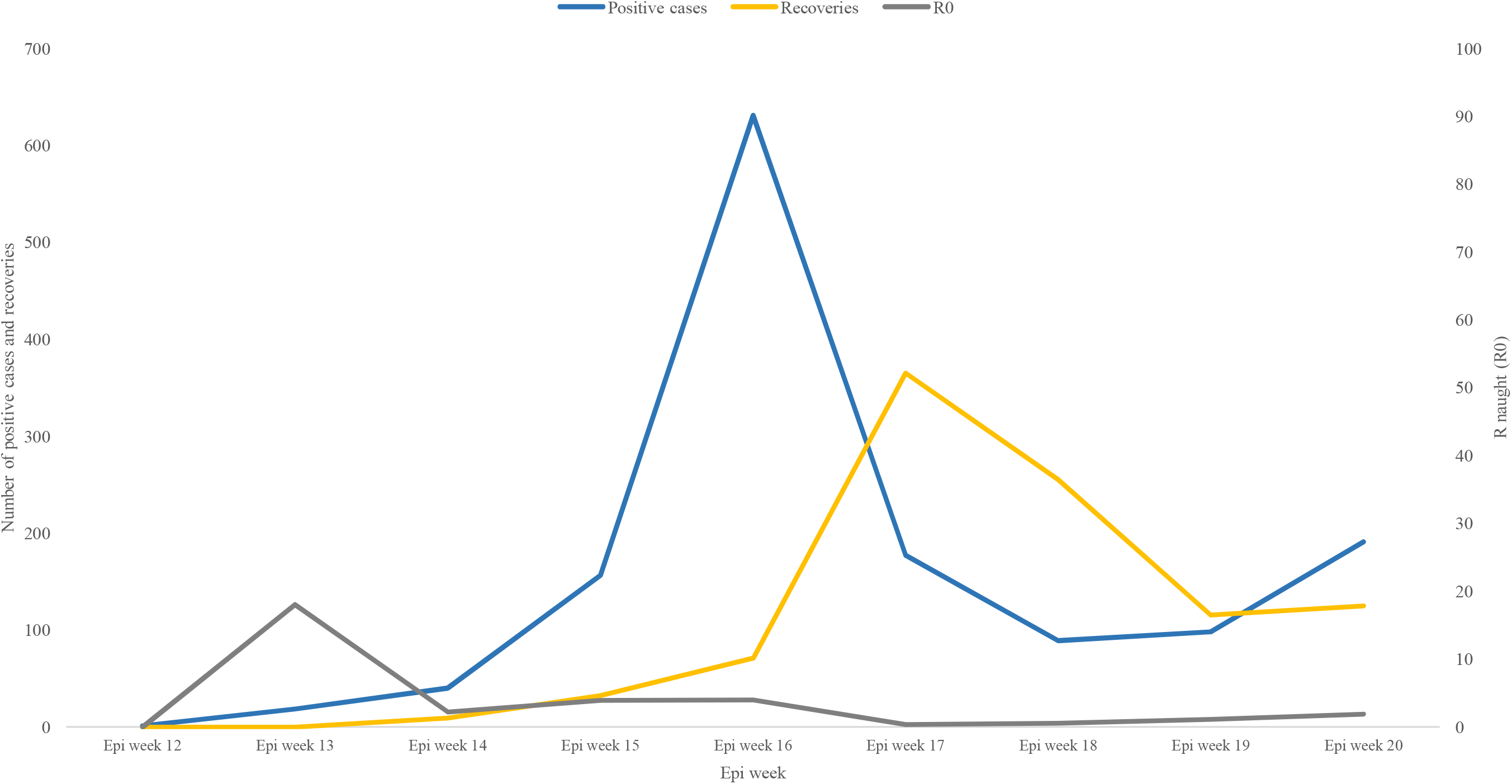
COVID-19 positive cases with the number of recoveries and R naught per epi week in Djibouti.

The figure also shows the progression of the basic reproduction number or the R naught (R0), which is an epidemiologic metric used to describe the contagiousness or transmissibility of infectious agents, by week. The highest reported R0 during the study period was during the epi week 13 (22-28 March 2020) with a value of 18. The weekly R naught varied during the study period from 0.3 to 18, with an overall R naught of 0.91 during the 9 weeks of the study.

Primary COVID-19 cases in Djibouti represented 2% (28) of the reported cases in the country during the study period. Secondary cases and contacts of the contacts represented 98% (1373) of the cases *(Figure 4)*. The low percentage of primary cases in Djibouti is explained by the closure of the land borders, port and airport on 18 March 2020. Therefore, no primary cases have been reported in the country after week 15 (5-11 April 2020).

**Figure 4.**
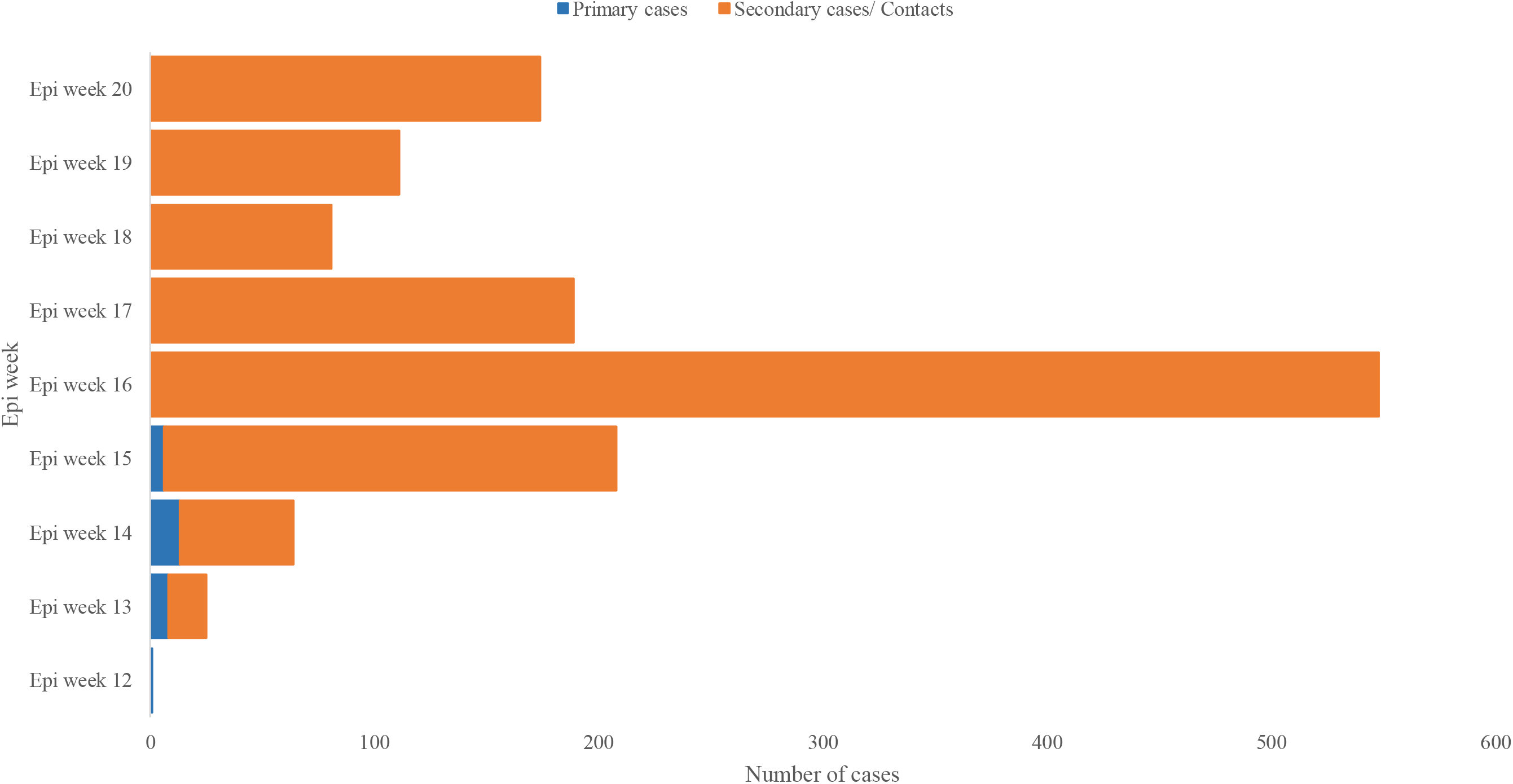
COVID-19 primary and secondary cases/ contacts reported in Djibouti, epi week 12 to epi week 20 /2020.

COVID-19 pandemic was reported in 4 out of 6 regions in the Republic of Djibouti. The affected regions were respectively: Djibouti-ville (the capital) with 83% of the reported cases, Ali Sabieh with 10% of the cases, Arta with 6% of the cases and Dikhil with only 1% of the cases reported during the study period *(Figure 5)*.

**Figure 5.**
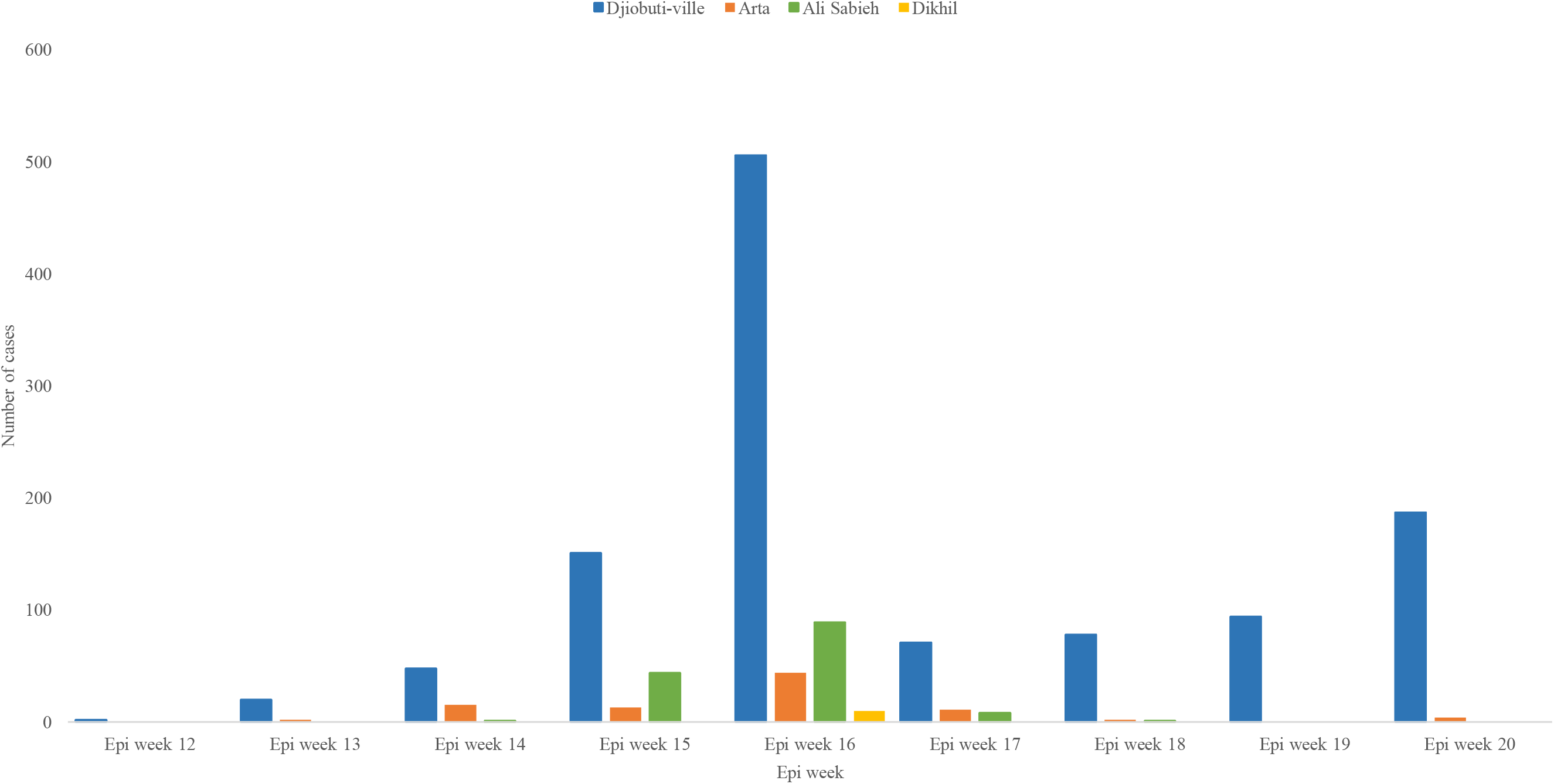
Distribution of COVID-19 cases reported in Djibouti by Region, epi week 12 to epi week 20, 2020.

## 4. Discussion

This is a descriptive study of the epidemiology of COVID-19 cases reported in Djibouti during the first two months of the detection of the virus in the country. It explains the response strategy put and adopted by the Ministry of Health of Djibouti together with its civilian and military partners during the same period, in close collaboration with WHO country office, which was followed strictly during the period of confinement announced by the government in the country between 23 March and 16 May 2020 (18).

The study findings show that the first peak of the COVID-19 pandemic in Djibouti started in the first week of April 2020 (epi week 15) and the number of cases started to decline during the third week of the month (epi week 18). Epi week 16 represented the top of the curve in Djibouti with 3,693 performed tests and with 631 positive cases detected, with a positivity rate of 17%. This is explained by the rise of the laboratory capacity to perform more tests per day, the detection of two clusters of COVID-19 in two hospitals during this period, one hospital in Djibouti-ville and the second in Ali Sabieh region (22), simultaneously with the start of the community transmission of the virus in the country.

The results of gender differences in the patients of COVID-19 in Djibouti are conforming with the results in other studies conducted in different countries and regions showing that the number of male patients affected by COVID-19 is almost the double of female patients (23, 24). Djibouti has a young population with a median age of 20 years old, and with 86.5% (832,067) of the population less than 49 years old according to the latest Djibouti Statistical Yearbook 2019 (12). This explains the high percentage of COVID-19 cases in the country, 73.2% (1025), below the age of 46 years old.

Djibouti showed a high percentage of recoveries among COVID-19 infected patients, 69% (972), during the study period. This was explained by the strict strategy of testing every suspected case of COVID-19 and contact tracing for all contacts. The patients were isolated at a very early stage of the infection, most of them asymptomatic, and treated according to the protocol agreed upon by the case management committee in the country. According to the case management protocol, Hydroxychloroquine and Azithromycin were introduced to all isolated patients, since day 1 of isolation up to they tested negative, with two consecutive swabs, with at least a period of 48 hours between the tests (16).

The mortality rate in Djibouti was one of the lowest in the region and globally, only 4 deaths (CFR: 0.3%). Deaths were elderly patients who were already in the Intensive Care Unit (ICU) even before being infected with COVID-19. They were recorded during the epi weeks: 15 (two deaths), 19 and 20 consecutively. Most of the cases of COVID-19 in Djibouti were contacts, 98% (1373), due to the strict interventions and decisions taken by the government of Djibouti of closing the airport, port and land borders, on 18 March 2020, just the following day of the detection of the first case in the country.

Djibouti-ville, the capital, was the most affected region in the country with COVID-19 cases, 83% (1157). This is due to the residence of more than 70-75% of the population in the capital according to the latest Djibouti Statistical Yearbook 2019 (12). Ali Sabieh comes as second region, with 10% (143), due to the outbreak of COVID-19 that was reported at the regional hospital in April 2020 (22).

It is important to mention that during the study period, according to the data from the Ministry of Health (MoH), 95-98% of the reported COVID-19 cases were completely asymptomatic and mainly detected through the contact tracing procedure. Therefore, only 2-5% of the cases has symptoms, mainly with very mild symptoms; therefore, it was not significant to register the clinical features of the patients of the newly emerging disease by the MoH, during the study period, in the country.

The strategy of response to the COVID-19 pandemic adopted by the government of Djibouti since the detection of the first case in the country up to the last day of the confinement, 16 May 2020, was based on four main pillars. These pillars were: 1) Testing every single COVID-19 suspected case according to the case definition, by taking the needed swabs for the PCR test; 2) Isolating all detected positive cases in a center of isolation prepared specifically for this reason by the government in Arta region; 3) Starting an early case management for all positive cases before even the development of symptoms; and 4) Conducting a strict contact tracing procedure for all contacts of the positive cases, testing them, and isolating those who were tested positive *(Figure 6)*.

**Figure 6.**
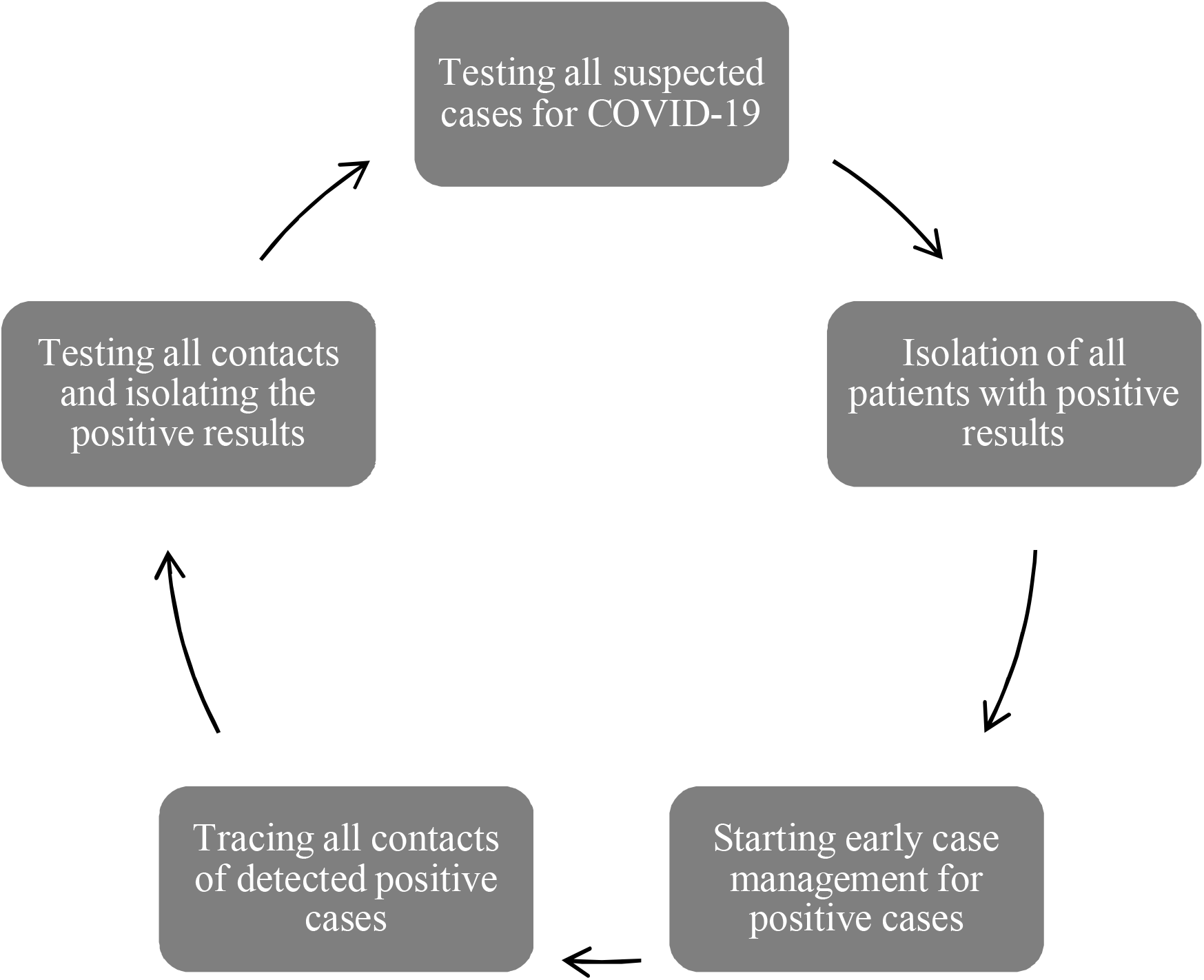
Cycle chart showing the 4 pillars on which the response strategy for COVID-19 in Djibouti was based during the study period.

This strategy permitted to protect Djibouti from having severe or critical cases of COVID-19 during the first two months of the circulation of the virus in the country. An objective that was targeted since the first day of the development of the preparedness and response plan due to the limited Intensive Care Unit (ICU) capacities in the country, as well as, the shortage in the human resources working for ICU.

Regarding the laboratory, Djibouti built its laboratory capacity for detection and confirmation of COVID-19 cases using the PCR machine in a very short and limited time, almost less than one week. The country started by the capacity for testing 100 samples per day and reached the capacity of testing up to 2,000 samples per day in the first week of April 2020. The government designated the laboratory of Hospital Bouffard, in the capital, as the only reference laboratory for the testing of COVID-19 cases.

During the study period, Djibouti conducted 17,532 tests for suspected COVID-19 cases as well as, the contacts detected through the contact tracing procedure, and the country was considered as a champion for COVID-19 tests in the African continent with the highest number of conducted tests per capita, 18.2 tests per 1000 habitant (17,532/ 962,451), up to 16 May 2020, considered higher than the triple of the average testing rates in the continent during the study period (25).

The early preparedness for the newly emerging disease; the transparency; the interventions taken by and the commitment of the government of Djibouti to save the lives of their people; lead to an adequate and an ideal response to the pandemic in the country. A close collaboration between MoH, WHO and other health sector partners during the preparation of the National Action Plan for preparedness and response to the COVID-19 in Djibouti, developed as a first draft in February 2020, reviewed and updated in March 2020; had its efficient and effective results during the response.

The preparedness and response plan for Djibouti was conforming with the WHO guidelines for surveillance strategies for COVID-19 human infection (20) published for the first time on 21 January 2020, with the latest update published on 10 May 2020. The response of the country was distinguished and a model among the countries at the horn of Africa, the WHO Eastern Mediterranean Region, and globally. It was also highly appreciated on the regional and global levels in many occasions (26).

The Ministry of Health of Djibouti understands that the coronavirus diseases (COVID-19) is still circulating in the country. Therefore, with the de-confinement started on 17 May 2020, a risk communication campaign was planned to sensitize the population to follow the barrier measures necessary to prevent a higher spread of the infection among the population. These measures include: 1) Frequent washing of hand with soap and water or an alcohol-based hand rub; 2) Preserving the physical distancing with others, either at home, at work or at any public place; and 3) Coughing or sneezing into a bended arm, or using a disposable tissue.

This campaign and other measures taken by the government of Djibouti will help the country to confront the disease. The vigilance of the MoH and the technical, operational and logistical support from WHO and other partners involved in the health sector in the country are the pillars to mitigate the harmful effects of the pandemic on the Djiboutian population.

## 5. Conclusion

Djibouti spent tremendous efforts to respond to the COVID-19 pandemic in the most adequate and efficient way, using the comparative advantages and harnessing the collaboration and collegiality between civilian and military health authorities that coordinated through the highest authorities in the country. The study described the epidemiological characteristics of the COVID-19 cases reported in the country and explained the response strategy followed by MoH and its partners. Conducting more studies is mandatory to deepen the understanding of the epidemiology of COVID-19, specifically, the clinical features of the emerging disease in the countries of the horn of Africa.

It is important to acknowledge that partnership, coordination, solidarity, vigilance, proactivity and commitment were essential to confront COVID-19 pandemic in Djibouti, as the capacities of the health system were only optimal when resources from military and civilian health apparatus were brought together and worked alongside WHO and other UN agencies, as well as, national and international partners.

## Data Availability

Yes, all data referred to in the manuscript are available.

## 8. Author contributions

All authors contributed equally to the manuscript.

## 9. Acknowledgements

The authors would like to thank His Excellency, President Ismail Omar Guelleh and the Government of Djibouti, for their high commitment by putting at disposal all the capacities of the country to respond to the COVID-19 pandemic, since the first day, in the most professional and efficient way. We would also like to thank the Djibouti Armed Forces, the Djiboutian National Gendarmerie and, the National Police of Djibouti, who contributed, in the most efficient way, to the response to the pandemic in the country. Finally, thanks to all medical and paramedical staff at the Ministry of Health and other entities; they are the real heroes of the response to the pandemic in the country.

